# Reliability and Concurrent Validity of a Computer Vision-Based Tool for Quantitative Finger Movement Analysis

**DOI:** 10.64898/2026.05.21.26353446

**Authors:** Arnav Maharshi, Bhakti Ladha, Rinkle Malani, Pallavi Palaskar

**Affiliations:** Independent Researcher and Developer of F.A.I.R. Chance©, a Computer Vision-based tool, Chhatrapati Sambhajinagar, Maharashtra, 431009, India; Postgraduate Student, Department of Neurophysiotherapy, MGM School of Physiotherapy, Chhatrapati Sambhajinagar, Maharashtra, 431003, a Constituent Unit of MGM Institute of Health Sciences, Navi Mumbai, India; Director, MGM School of Physiotherapy, Chhatrapati Sambhajinagar, Maharashtra, 431003, a Constituent Unit of MGM Institute of Health Sciences, Navi Mumbai, India; Professor and Head of Department of Neurophysiotherapy, MGM School of Physiotherapy, Chhatrapati Sambhajinagar, Maharashtra, 431003, a Constituent Unit of MGM Institute of Health Sciences, Navi Mumbai, India

**Keywords:** artificial intelligence, assistive technology, computer vision, fine motor evaluation, hand function

## Abstract

**Background:** Accurate evaluation of fine motor abilities is a key aspect of neurological rehabilitation. However, conventional approaches like goniometry are limited by variations among raters and their difficulty in detecting active movement. On the other hand, computer vision-based software delivers non-invasive and quantitative analysis of hand movements. An innovative computer-vision-based software tool, F.A.I.R. Chance©, was developed to track and analyze individual finger joint movements on a camera-equipped laptop and give real-time numerical feedback. However, its metrics require validation in a healthy population before the tool can be used for clinical purposes.

**Objective:** To assess the reliability and validity of finger movement assessment by the F.A.I.R. Chance computer vision-based tool in healthy adult participants.

**Methods:** An observational cross-sectional study was done at MGM School of Physiotherapy, comprising 30 healthy participants between 18 and 60 years of age. Finger movements like flexion, extension, abduction, and adduction were measured with a standard handheld goniometer. These same finger movements were then measured with the tool at two time points separated by a 30-minute interval to determine the test-retest reliability. The tool’s measurements were compared with the goniometric measurements to determine its concurrent validity. Test-retest reliability was checked by the Intra-class Correlation Coefficient ICC (2,1), while concurrent validity was tested through Pearson’s correlation coefficients.

**Results:** Metacarpophalangeal and proximal interphalangeal joint motions demonstrated moderate to good test-retest reliability (ICC: 0.716–0.953) for the F.A.I.R. Chance tool. However, distal interphalangeal joint movements had lower consistency. Good reliability (ICC: 0.754–0.908) was seen for movements of abduction and adduction in the fingers. Strong concurrent validity for extension movements of the metacarpophalangeal joints (r=0.760–0.914) and moderate concurrent validity for flexion movements of the metacarpophalangeal joints (r=0.427–0.604) was demonstrated for all fingers for the F.A.I.R. Chance tool. Concurrent validity for adduction and abduction movements demonstrated a low to fair correlation with goniometric measurements (r=0.210–0.440). This is consistent with previous research showing poor agreement between goniometry and adduction-abduction movements of the fingers

**Conclusion:** The F.A.I.R. Chance tool shows good reliability and acceptable concurrent validity to assess fine motor movements in the healthy adult population. This sets a basis for further clinical study of the tool in the target population with fine motor impairments.

## Introduction

Fine motor activity includes the precise, coordinated activity of the small muscles of the hands and digits, allowing tasks of everyday life such as writing, dining, buttoning, and handling objects. Limitations in fine motor skills are noted in neurological conditions like stroke, cerebral palsy, Parkinson’s disease, multiple sclerosis, traumatic brain injury, and spinal cord injury, leading to impairments in task-based performance and quality of life.

Conventional clinical tools like the Nine-Hole Peg Test, Purdue Pegboard Test, Jebsen-Taylor Hand Function Test, and Box and Block Test deliver specific task gains, providing less information on the level of joint motion ^[1-4]^. While conventional goniometry is used in daily practice, it is prone to inter- and intra-rater variations, lacking the capability to determine the active movement patterns ^[5-7]^. These limitations underscore the need for objective, reliable, and real-time assessment methods enabled by recent technological advances.

While wearable sensors have shown promise in rehabilitation monitoring ^[8, 9]^, recent innovations in visual AI facilitate non-invasive, image- and video-based motion analysis that records fine movement patterns independent of wearable technology ^[10,11]^. Prior to this study, the F.A.I.R. (Fine-motor Artificial Intelligence-assisted Rehabilitation) Chance© tool was developed to leverage computer vision and hand-tracking algorithms (using Google MediaPipe framework) to detect and analyze fine motor movements in real time using a single camera-equipped device (smartphone/laptop/tablet). The system detects and tracks 21 hand-knuckle joints, evaluates joint angles, ranges of motion, and movement patterns relevant to functional rehabilitation (e.g., flexion/extension, abduction/adduction, thumb opposition), and provides immediate, quantitative feedback to patients and therapists, all in real-time. The data offered can be used for progress monitoring, adaptive goal-setting, and patient motivation toward recovery goals. However, its metrics require validation before using it for clinical purposes.

Therefore, the aim of this study was to assess the test-retest reliability of the F.A.I.R. Chance tool and the objective was to quantify its concurrent validity relative to conventional goniometry.

## Methods

### Design

This study employed an observational cross-sectional design.

### Sample size

Thirty healthy adult participants were recruited for this study. The chosen sample size of 30 participants satisfies the minimum requirement for statistical analysis of reliability coefficients such as Intraclass Correlation Coefficient (ICC), Cronbach’s alpha, and concurrent validity measures, including Pearson’s or Spearman’s correlation analysis. Additionally, a sample size of 30 is generally considered adequate to approximate normal distribution according to the Central Limit Theorem, thereby allowing meaningful parametric statistical analysis.

### Duration and place of study

The study was conducted over three months from December 2025 to February 2026 at the Department of Neuro-physiotherapy, MGM School of Physiotherapy, Chhatrapati Sambhajinagar, Maharashtra, India.

### Eligibility Criteria

Participants with the age range of 18 to 60 years with no previous history of any neurological disorders; no musculoskeletal disorders that can affect hand function; a normal range of motion and strength in both hands and the ability to follow instructions were included in the study. Participants with visual impairment resulting in difficulty performing the task or those who refused to participate were excluded from the study.

### Procedure

Participants were given all the necessary information and details of the procedure regarding this study. Basic information like age, gender, and hand dominance was obtained from the participants.

Participants were asked to sit in a comfortable position (Fig. 1) and later asked to perform flexion, extension, adduction, and abduction of all four fingers (index, middle, ring, and little finger). First, assessor 1 measured the flexion-extension movements at the metacarpophalangeal (MCP), proximal interphalangeal (PIP), and distal interphalangeal (DIP) joints for all four fingers using the standard goniometer. Similarly, adduction-abduction movements of all the fingers were measured using a standard goniometer. Then, the participants performed these same finger movements in front of a camera-equipped laptop running the F.A.I.R. Chance tool that tracked and evaluated these movements, which were recorded by assessor 2 (time point 1). This tool measured the flexion and extension movements for the metacarpophalangeal (MCP), proximal interphalangeal (PIP), and distal interphalangeal (DIP) joints for all four fingers by measuring the angle formed at the respective joint (Fig. 2 and Fig. 3). The tool also measured the adduction-abduction of fingers relative to the midline axis (middle finger) of the hand. The measurements were retaken on the F.A.I.R. Chance tool after a 30-minute interval by the same assessor 2 (time point 2). A 30-minute interval between test sessions was selected to prevent neuromuscular fatigue while minimizing physiological fluctuations in joint ROM. The movements assessed were simple finger actions in healthy adults, with no meaningful learning curve or diurnal variation within this interval. This interval was deemed sufficient to minimise practice effects and recall bias while avoiding the potential confounding biological variability introduced by longer inter-session gaps. Assessor 2 was blinded to the readings of goniometric measurements.

### Statistical Analysis

Test-retest reliability was calculated using the Intra-class Correlation Coefficient ICC (2,1) with 95% confidence interval. Koo and Li guideline ^[23]^ was used for interpreting the Intra-class Correlation Coefficients values. Concurrent validity was assessed with Pearson’s correlation coefficients. The classification proposed by Schober et al. (2018) ^[24]^ was used to interpret Pearson’s correlation coefficients. SPSS software version 25.0 was used for the analysis.

### Ethical Considerations

This study was approved by the Institutional Ethics Committee (ECRHS) of MGM Medical College and Hospital, Chhatrapati Sambhajinagar, Maharashtra, India (Reference number: MGM-ECRHS/2025/35). Written informed consent was obtained from all the participants before their enrollment in this study, and participants were informed that data collected would be published while maintaining confidentiality.

## Results

### 1. Demography

Thirty participants were included in this study; averaging 28.6 ± 10.5 years; among these 66.7% were female participants. 22 of the 30 participants (73.3%) were right-hand dominant, whereas 8 out of the 30 participants (26.7%) were left-hand dominant.

**Table 1.** Characteristics of the participants included in this study (n=30)

| Characteristics | n | Percentage/Mean ± SD |
| --- | --- | --- |
| Total participants | 30 | 100.0% |
| Age (years) | | $28.6 \pm 10.5$ (range: 18–58) |
| Gender |  |  |
| Male | 10 | 33.3% |
| Female | 20 | 66.7% |
| Hand Dominance |  |  |
| Right | 22 | 73.3% |
| Left | 8 | 26.7% |

### 2. Test-Retest Reliability

Test-retest analysis showed good and acceptable intra-class correlation coefficients for almost all of the metacarpophalangeal and proximal interphalangeal joint actions. On the other hand, actions at the distal interphalangeal joints showed somewhat reduced reliability. The highest reliability was seen for the extension movement of the MCP joint. Reliability for the abduction movement was found to be superior in comparison to the adduction movement for all fingers.

**Table 2.** Values for test-retest reliability of the F.A.I.R. Chance tool: Intraclass Correlation Coefficient (ICC) for all fingers and joints for flexion and extension.

|  | INDEX FINGER |  |  |  |  |  |
| --- | --- | --- | --- | --- | --- | --- |
|  | MCP |  | PIP |  | DIP |  |
|  | Flexion | Extension | Flexion | Extension | Flexion | Extension |
| Intraclass Correlation Coefficient (ICC) | 0.716<br>p<0.0001 | 0.916<br>p<0.0001 | 0.391<br>p=0.002 | 0.556<br>p<0.0001 | 0.611<br>p<0.0001 | 0.513<br>p=0.003 |
| MIDDLE FINGER |  |  |  |  |  |  |
|  | MCP |  | PIP |  | DIP |  |
|  | Flexion | Extension | Flexion | Extension | Flexion | Extension |
| Intraclass Correlation Coefficient (ICC) | 0.612<br>p<0.0001 | 0.953<br>p<0.0001 | 0.815<br>p<0.0001 | 0.795<br>p<0.0001 | 0.883<br>p<0.0001 | 0.467<br>p=0.001 |
| RING FINGER |  |  |  |  |  |  |
|  | MCP |  | PIP |  | DIP |  |
|  | Flexion | Extension | Flexion | Extension | Flexion | Extension |
| Intraclass Correlation Coefficient (ICC) | 0.909<br>p<0.0001 | 0.896<br>p<0.0001 | 0.761<br>p<0.0001 | 0.826<br>p<0.0001 | 0.883<br>p<0.0001 | 0.893<br>p<0.0001 |
| LITTLE FINGER |  |  |  |  |  |  |
|  | MCP |  | PIP |  | DIP |  |
|  | Flexion | Extension | Flexion | Extension | Flexion | Extension |
| Intraclass Correlation Coefficient (ICC) | 0.862<br>p<0.0001 | 0.901<br>p<0.0001 | 0.817<br>p<0.0001 | 0.817<br>p<0.0001 | 0.812<br>p<0.0001 | 0.475<br>p<0.0001 |

**Table 3.** Interpretation* of the values for test-retest reliability of the F.A.I.R. Chance tool for all fingers and joints for flexion and extension.

| Finger | MCP Joint (ICC) | PIP Joint (ICC) | DIP Joint (ICC) | Notes |
| --- | --- | --- | --- | --- |
| **Index** | Extension:<br>Excellent (>0.9)<br>Flexion:<br>Moderate (0.716) | Extension:<br>Moderate (0.556)<br>Flexion:<br>Poor (0.391) | Extension:<br>Moderate (0.513)<br>Flexion:<br>Moderate (0.611) | Lower value for DIP due to tracking challenges |
| **Middle** | Extension:<br>Excellent (>0.9)<br>Flexion: Moderate (0.612) | Extension:<br>Good (0.795)<br>Flexion:<br>Good (0.815) | Extension:<br>Poor (0.467)<br>Flexion:<br>Good (0.883) | Proximal joints strongest |
| **Ring** | Extension:<br>Good (0.896)<br>Flexion:<br>Excellent (0.909) | Extension: Good (0.826)<br>Flexion: Good (0.761) | Extension: Good (0.893)<br>Flexion: Good (0.883) | Highest overall reliability; best visibility |
| **Little** | Extension:<br>Excellent (0.901)<br>Flexion: Good (0.862) | Extension:<br>Good (0.817)<br>Flexion:<br>Good (0.817) | Extension: Poor (0.475)<br>Flexion: Good (0.812) | Affected by size and overlap |

**Table 4.**
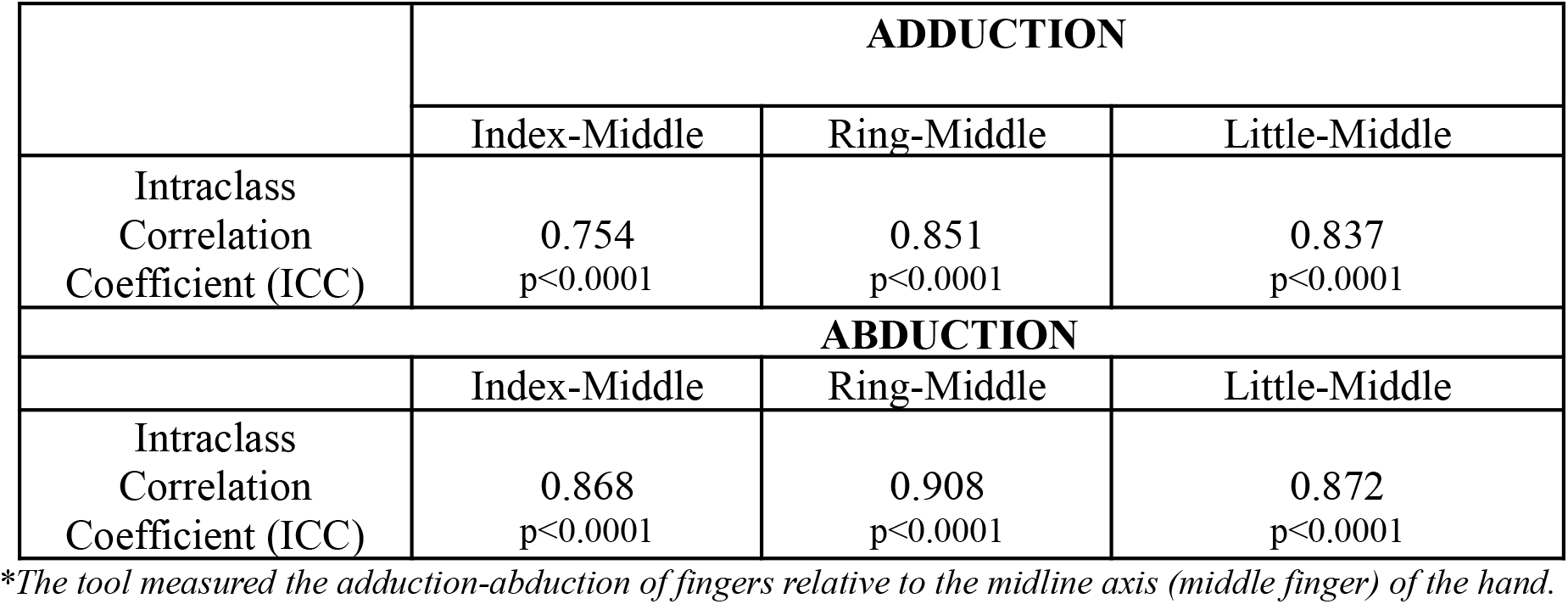
Values for test-retest Reliability of the F.A.I.R. Chance tool: Intraclass Correlation Coefficient (ICC) for all fingers for abduction and adduction.

|  | ADDUCTION |  |  |
| --- | --- | --- | --- |
|  | Index-Middle | Ring-Middle | Little-Middle |
| Intraclass Correlation Coefficient (ICC) | 0.754<br>p<0.0001 | 0.851<br>p<0.0001 | 0.837<br>p<0.0001 |
|  | ABDUCTION |  |  |
|  | Index-Middle | Ring-Middle | Little-Middle |
| Intraclass Correlation Coefficient (ICC) | 0.868<br>p<0.0001 | 0.908<br>p<0.0001 | 0.872<br>p<0.0001 |

*\*The tool measured the adduction-abduction of fingers relative to the midline axis (middle finger) of the hand.*

**Table 5.** Interpretation* of the values for test-retest Reliability of the F.A.I.R. Chance tool across all fingers for abduction and adduction.

| Movement | ICC Range | Interpretation |
| --- | --- | --- |
| **Adduction** | 0.754–0.851 | Good |
| **Abduction** | 0.868–0.908 | Good to Excellent |

*\*Interpretation of Intraclass Correlation Coefficient (ICC) values (Koo and Li guideline) <sup>[23]</sup>*

| Reliability coefficient value | Interpretation |
| --- | --- |
| 0.90 and up | Excellent |
| 0.75–0.9 | Good |
| 0.5–0.75 | Moderate |
| below 0.50 | Poor |

### 3. Concurrent Validity

Strong to very strong correlation for the extension movement for all fingers at the MCP joint was seen between the F.A.I.R. Chance tool and goniometric measurement when analysis for the concurrent validity was performed. Moderate correlation was seen for the flexion movement at the MCP joint, whereas low to moderate correlation was noted for the PIP and DIP joint motions. Concurrent validity for extension motion was greater for the metacarpophalangeal joints than for the PIP and DIP joints.

**Table 6.**
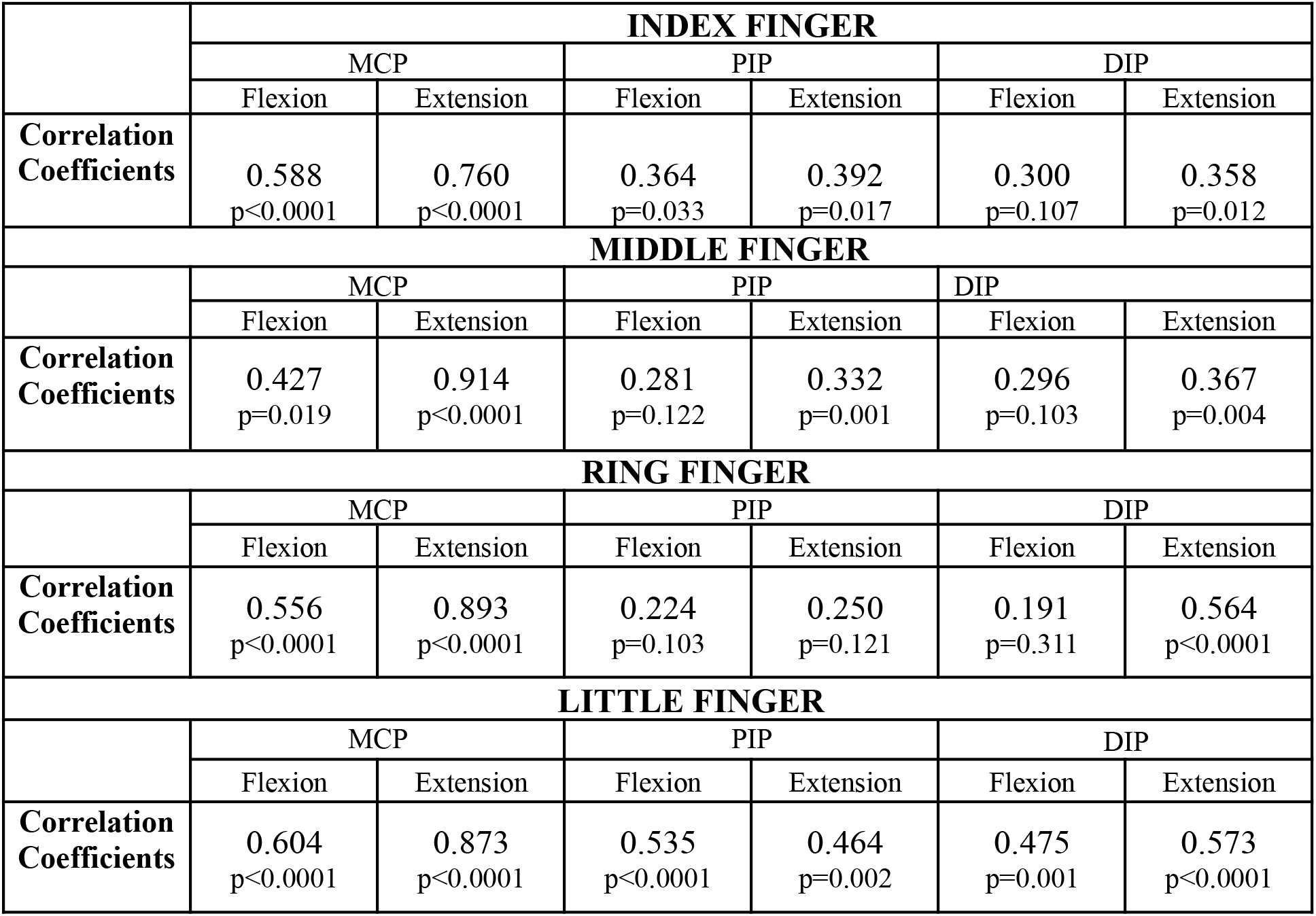
Concurrent Validity: Pearson’s coefficient for all fingers and joints for flexion and extension.

**Table 7.** Interpretation of Concurrent Validity: Pearson’s coefficient for all fingers and joints for flexion and extension.

| Finger | MCP Joint (r) | PIP Joint (r) | DIP Joint (r) | Notes |
| --- | --- | --- | --- | --- |
| Index | Extension:<br>Strong-Very strong<br>(0.760–0.914)<br>Flexion: Moderate<br>(0.5–0.7) | Flexion &<br>Extension<br>Weak (0.3) | Flexion &<br>Extension<br>Weak (0.3) | Strongest for MCP<br>joint extension |
| Middle | Extension:<br>Strong-Very strong<br>(0.760–0.914)<br>Flexion: Moderate<br>(0.5–0.7) | Flexion &<br>Extension<br>Weak (0.3) | Flexion &<br>Extension<br>Weak (0.3) | Aligns with<br>proximal joint<br>advantages. |
| Ring | Extension:<br>Strong-Very strong<br>(0.760–0.914)<br>Flexion:<br>Moderate-good<br>(0.5–0.76) | Flexion &<br>Extension<br>Weak (0.3) | Extension:<br>Moderate (0.564)<br>Flexion: Weak<br>(0.191) | Consistent with high<br>reliability here. |
| Little Finger | Extension:<br>Strong-Very strong<br>(0.7–0.914)<br>Flexion: Moderate<br>(0.5–0.7) | Flexion &<br>Extension<br>Moderate<br>(0.535–0.464) | Flexion &<br>Extension<br>Moderate<br>(0.475–0.573) | Limited by<br>anatomical<br>challenges. |

**Table 8.** Concurrent Validity: Pearson’s coefficient for all fingers for adduction and Abduction.

|  | ADDUCTION |  |  |
| --- | --- | --- | --- |
|  | Index-Middle | Ring-Middle | Little-Middle |
| Correlation Coefficients | 0.366<br>p<0.0001 | 0.210<br>p=0.109 | 0.384<br>p=0.001 |
|  | ABDUCTION |  |  |
|  | Index-Middle | Ring-Middle | Little-Middle |
| Correlation Coefficients | 0.440<br>p<0.0001 | 0.285<br>p=0.033 | 0.272<br>p=0.041 |

*\*The tool measured the adduction-abduction of fingers relative to the midline axis (middle finger) of the hand.*

**Table 9.** Interpretation* of Concurrent Validity for Abduction and Adduction for all fingers.

| **Movement** | **r Range** | **Interpretation** |
| --- | --- | --- |
| Abduction | 0.272–0.440 | Weak |
| Adduction | 0.210–0.384 | Weak-moderate |

\*Schober et al. (2018) <sup>[24]</sup> Interpretation

| **Pearson's r Category** | **Interpretation** |
| --- | --- |
| 0.00–0.10 | Negligible |
| 0.10–0.39 | Weak |
| 0.40–0.69 | Moderate |
| 0.70–0.89 | Strong |
| 0.90–1.0 | Very strong |

## Discussion

### 1. Demography

This study analyzed reliability and concurrent validity by using F.A.I.R. (Fine Motor Artificial Intelligence-Assisted Rehabilitation) Chance tool, a computer vision-based setup made for the objective testing of finger actions. To evaluate these features in a healthy population is an important step before clinical use, mainly for therapy, where specific details of hand and finger motions are a basic requirement for assessment, goal-setting, and monitoring of post-therapy outcomes ^[12-15]^.

Young adults were commonly selected in initial assessment studies because they have neuromuscular coordination, show enough range of motion, and less age-related structural deformities. This homogeneity limits variability and allows clear assessment of measurement-based performance and precision ^[16]^. The same age range population has been seen in previous studies that targeted hand actions and an AI-based movement tracking set-up that supported the suitability of this current sample in research designs ^[18, 19]^.

Sex-related variations were unlikely to have affected the reliability or validity of the results. While less variation in hypermobility and finger motion between both genders was seen, these differences did not appear to affect the properties of joint angle measurement ^[18]^. Therefore, the characteristics of all participants in this study are unlikely to have formed any sort of bias in the outcomes.

### 2. Test-retest reliability

The F.A.I.R. Chance tool showed good test-retest reliability in most finger actions, with particularly higher ICC values at the metacarpophalangeal (MCP) and proximal interphalangeal (PIP) joints. These data indicate that the tool is stable and reliable over several assessments, which is a key necessity for clinical use and research purposes ^[18]^.

The reliability of the index finger changes on the basis of the joint test and motion performed. Extension of the MCP joint indicated strong reliability, whereas flexion movement of the PIP and DIP joints demonstrated somewhat lower ICC values ^[18, 19]^. This finding agrees with earlier literature that focused on the challenges of correctly measuring actions of distal finger joints, due to their movement limitations, body structures, and a greater risk of blocking sight during assessment of movements ^[16]^.

This type of similarity was observed for the middle finger. Extension at the MCP joint demonstrated excellent reliability, while movements of the PIP had good reliability, but extension at the DIP had reduced consistency. Similar findings were reported in previous studies that had employed optical motion trackers and sensor-based technologies, where distal joints faced challenges due to their limited ranges and the difficulty in detecting their anatomical points using camera-based trackers ^[18]^.

The ring finger showed the most reliable results for MCP, PIP, and DIP ^[18, 19]^. Similar findings were noted before for computer-based motion monitoring studies, further supporting the accuracy of these results ^[18, 19]^. Overall, the results revealed that the use of the F.A.I.R. Chance tool could be clinically applicable for assessing the movement of the ring finger.

The little finger showed good reliability for most joints, while extension at the DIP was seen with lesser ICC values. Due to its smaller size, reduced mobility, and overlapping with nearby fingers, it was difficult to assess it correctly. This may have affected both manual and computer-based techniques. Still, the general reliability for little fingers can be considered in normal ranges for both clinical use and research studies ^[18]^.

The F.A.I.R. Chance tool showed good reliability for adduction and abduction actions. This is of great importance as adduction-abduction movements of fingers are difficult to measure with a traditional goniometer due to misalignment and increased dependence on the examiner for measurement. The high reliability achieved indicates that AI-based methods can reduce many issues ^[18, 19]^.

Abduction movements proved superior to movements of adduction in terms of reliability. This may be due to clear visual spacing between fingers during abduction movement, which allows AI to detect anatomical tracking landmarks more accurately. Inter-finger gaps were decreased in adduction, which resulted in increased overlapping. Thus, ICC values were higher for abduction due to better inter-finger spacing. Even so, overall reliability for adduction-abduction movements of fingers demonstrates the tool’s potential to detect finger actions that are otherwise poorly evaluated in a regular clinical setting.

### 3. Concurrent validity

Concurrent validity showed a strong to very strong correlation between the F.A.I.R. Chance tool and goniometric measurements for extension at all MCP joints. Flexion at the MCP joint showed moderate correlations, while PIP and DIP joint range of motion showed low to moderate correlations. These findings align with previous research on movement tracking technology, which demonstrated higher agreement with goniometry for greater ranges and more prominent anatomical references ^[16, 17]^.

Greater concurrent validity was noted for the extension of the MCP joint, which indicated that the F.A.I.R. Chance tool is effective in tracking finger movements with an increased range of motion. On the other hand, the smaller correlations at the PIP and DIP joints showed basic limitations with manual goniometry, due to reduced reliability when assessing small joints. These differences should not be interpreted as a limitation of the tool, but rather as a reflection of the inherent imprecision of the reference standard used for comparison.

Concurrent validity of the F.A.I.R. Chance tool for adduction and abduction motions ranged from low to fair. This is consistent with previous research showing poor agreement between goniometry and adduction-abduction movements of the fingers ^[22]^. Significantly, in spite of the lower validity correlations for adduction and abduction movements, the F.A.I.R. Chance tool had strong test-retest reliability ^[18-20]^.

This inconsistency is the key challenge in testing assistive technologies: lower r values do not necessarily indicate tool inaccuracy when the reference standard used for comparison has known measurement limitations. This suggests that the AI-based tool may yield a more stable and objective approach to analyze finger movements when compared to conventional manual techniques ^[18-21]^.

## Clinical Implications

The findings suggest that the F.A.I.R. Chance tool is a reliable and valid tool for objective assessment of fine motor movements in healthy adults. Its non-invasive nature, together with joint-specific quantitative results in real-time, provides advantages over conventional assessment approaches. The tool appears suitable for assessing adduction-abduction finger actions and MCP joint movements, which are clinically notable but otherwise hard to assess accurately by conventional methods.

The tool offers a distinct clinical advantage in reducing the subjective variability in measuring the deficit in finger movements and the subsequent improvement. Instead, clear numerical data are available in real time to assess the individual finger joint movements. Establishing these measurement characteristics in a healthy population provides a strong basis for future studies to include neurologically ill patients, where fine motor impairments are more prevalent, severe, and clinically meaningful. Repeated, standardized assessments will be required to monitor recovery and to measure the impact of rehabilitative approaches.

Effective pincer movements and cupping movements of the fingers are necessary for writing, picking up objects, or eating with fingers. These movements require a complex extensor-flexor synergy, most notably the ability to maintain extension at PIP and DIP joints while regulating MCP flexion. Objective measurement of MCP joint extension is therefore an important metric for tracking the recovery trajectory in paralyzed individuals. The F.A.I.R. Chance tool demonstrates strong reliability in tracking the MCP joint extension in healthy individuals, suggesting robust clinical applicability of the tool.

Opposition of thumb, an equally crucial movement for functional fine motor movements, is affected in cases of stroke, cerebral palsy, and other neurological disorders. Beyond the assessment of finger movements in this study, the tool is capable of providing numerical feedback for opposition movements of the thumb with each finger by measuring the inter-tip distance. However, this feature could not be validated in the current healthy population of this study due to the absence of fine motor deficits, but it provides a foundation for future clinical validation in affected individuals.

## Limitations

1. The reliability of the tool for the opposition movement of the thumb could not be assessed due to a ceiling effect, as all healthy participants demonstrated complete opposition movements at both time points. Hence, there was no measurable variation between the two sessions on which ICC could be meaningfully computed. Similarly, concurrent validity could not be assessed, as the conventional goniometer is not designed to measure opposition in this manner.
2. Potentially, the tool is able to measure the deficit or improvement in finger movements in increments of 1–2%. However, since the study was conducted in a healthy population, the minimum detectable change (MDC) could not be determined and remains to be established in clinical populations.

## Conclusion

The F.A.I.R. Chance tool is a robust, reliable, and validated computer vision-based tool for the accurate evaluation of finger joint movements in healthy adults. Future validation studies in neurological populations, particularly in patients with stroke and cerebral palsy, are necessary to establish clinical applicability.

## Supporting information

STROBE Checklist

## Data Availability

All data are available from the corresponding author on reasonable request.

## Data Availability Statement

Data are available from the corresponding author on reasonable request.

