## Supplementary material for "Reliability and Concurrent Validity of a Computer Vision-Based Tool for Quantitative Finger Movement Analysis": STROBE Checklist

### STROBE Checklist- Cross-Sectional Study

*Reporting Guideline: STROBE Statement: von Elm E et al., PLoS Med. 2007;4(10):e296.*

| No. | Section / Topic | Checklist Item | Location in Manuscript |
|---|---|---|---|
| **TITLE & ABSTRACT** | | | |
| 1 | Title and abstract | (a) Indicate the study's design with a commonly used term in the title or the abstract<br>(b) Provide in the abstract an informative and balanced summary of what was done and what was found | (a) Study design described as 'observational cross-sectional' in Methods. Title indicates 'Computer Vision-Based Tool' and measurement study context.<br>(b) Structured abstract on p.1-2 covers background, objective, methods (design, participants, measurements), results (ICC and r values by joint), and conclusion. |
| **INTRODUCTION** | | | |
| 2 | Background / rationale | Explain the scientific background and rationale for the investigation being reported | Introduction, p.2: Limitations of conventional goniometry (inter/intra-rater variability, inability to detect active movement) explained. Wearable sensors and computer vision tools contextualised. F.A.I.R. Chance tool development and need for validation stated. |
| 3 | Objectives | State specific objectives, including any prespecified hypotheses | Introduction, p.3: Aim stated: to assess test-retest reliability of F.A.I.R. Chance. Objective stated: to quantify concurrent validity relative to conventional goniometry. |
| **METHODS** | | | |
| 4 | Study design | Present key elements of study design early in the paper | Methods- Design, p.3: 'This study employed an observational cross-sectional design.' |
| 5 | Setting | Describe the setting, locations, and relevant dates, including periods of recruitment, exposure, follow-up, and data collection | Methods- Duration and place of study, p.3: Department of Neuro-physiotherapy, MGM School of Physiotherapy, Chhatrapati Sambhajinagar, Maharashtra, India. Study period: December 2025–February 2026 (3 months). |
| 6 | Participants | (a) Give the eligibility criteria, and the sources and methods of selection of participants<br>(b) For matched studies, give matching criteria and number matched and unmatched | (a) Eligibility Criteria, p.3: Inclusion- age 18-60, no neurological or musculoskeletal disorders, normal ROM and strength, ability to follow instructions. Exclusion- visual impairment affecting task performance, refusal to participate.<br>(b) Not applicable (no matched design). |
| 7 | Variables | Clearly define all outcomes, exposures, predictors, potential confounders, and effect modifiers. Give diagnostic criteria if applicable | Methods- Procedure, p.3-4: Primary outcomes- ICC for test-retest reliability, Pearson's r for concurrent validity. Movements assessed: flexion, extension, abduction, adduction at MCP, PIP, and DIP joints for all four fingers (index, middle, ring, little). Figures 1-3 (p.4) illustrate setup and tool interface. |

| No. | Section / Topic | Checklist Item | Location in Manuscript |
|---|---|---|---|
| 8 | Data sources / measurement | For each variable of interest, give sources of data and details of methods of assessment. Describe comparability if more than one assessment method is used | Methods- Procedure, p.3-4: Assessor 1 measured goniometric ROM. Assessor 2 (blinded to goniometry) recorded F.A.I.R. Chance tool measurements at two time points 30 minutes apart. Tool measures joint angles via Google MediaPipe (21 hand-knuckle landmarks); adduction-abduction measured relative to midline (middle finger axis). Fig. 2-3 (p.4) show measurement interface. |
| 9 | Bias | Describe any efforts to address potential sources of bias | Methods: Procedure, p.3-4: Assessor 2 blinded to goniometric readings. Standardised seating position (Fig. 1, p.4). 30-minute interval selected to prevent neuromuscular fatigue, minimise practice effects, recall bias, and diurnal biological variation. |
| 10 | Study size | Explain how the study size was arrived at | Methods: Sample size, p.3: n=30 justified as meeting minimum requirement for ICC, Cronbach's alpha, and correlation analyses. Sample of 30 satisfies Central Limit Theorem for parametric analysis. No formal a priori power calculation reported. |
| 11 | Quantitative variables | Explain how quantitative variables were handled in the analyses. If applicable, describe which groupings were chosen and why | Methods: Statistical Analysis, p.5: ICC and Pearson's r treated as continuous. ICC interpreted by Koo and Li (2016) categories (Poor <0.5, Moderate 0.5–0.75, Good 0.75–0.9, Excellent ≥0.9). Pearson's r interpreted by Schober et al. (2018) categories (Negligible, Weak, Moderate, Strong, Very strong). |
| 12 | Statistical methods | (a) Describe all statistical methods, including those used to control for confounding<br>(b) Describe methods for subgroup analyses<br>(c) Explain how missing data were addressed<br>(d) Describe analytical methods accounting for sampling strategy<br>(e) Describe any sensitivity analyses | (a) ICC(2,1) with 95% CI (test-retest reliability); Pearson's r (concurrent validity). SPSS v25.0. p.5.<br>(b) Analysis stratified by finger (4), joint (3), and movement type- reported in Tables 2-9 (p.6-10).<br>(c) Missing data not discussed; no missing data apparent from results.<br>(d) Not applicable; convenience sample of healthy adults in a single setting.<br>(e) No sensitivity analyses conducted or reported. |
| **RESULTS** | | | |
| 13 | Participants | (a) Report numbers of individuals at each stage of study- e.g., potentially eligible, examined, confirmed eligible, included, completing follow-up, and analysed<br>(b) Give reasons for non-participation at each stage<br>(c) Consider use of a flow diagram | (a) 30 participants enrolled and analysed. Results- Demography, p.5; Table 1, p.5.<br>(b) Reasons for non-participation not reported.<br>(c) No participant flow diagram included. |
| 14 | Descriptive data | (a) Give characteristics of study participants and information on exposures and potential confounders<br>(b) Indicate number of participants with missing data for each variable of interest | (a) Table 1, p.5: Age (mean 28.6 ± 10.5 years, range 18-58), sex (33.3% male, 66.7% female), hand dominance (73.3% right, 26.7% left).<br>(b) No missing data reported for any variable. |
| 15 | Outcome data | Report numbers of outcome events or summary measures | Tables 2-5 (p.6-7): ICC values with p-values for all 24 flexion/extension joint-finger combinations and 6 abduction/adduction pairs. Tables 6-9 (p.8-10): Pearson's r with p-values for the same combinations. |
| 16 | Main results | (a) Give unadjusted estimates and precision (e.g., 95% CI) | (a) ICC values with 95% CI and p-values; Pearson's r with p-values reported. Tables 2, 4, 6, 8 (p.6-9). |

| No. | Section / Topic | Checklist Item | Location in Manuscript |
|---|---|---|---|
| | | (b) Report category boundaries when continuous variables were categorised<br>(c) If relevant, consider translating estimates into absolute risk | (b) ICC and r interpretive categories applied in Tables 3, 5, 7, 9 (p.7, 9, 10).<br>(c) Not applicable. |
| 17 | Other analyses | Report other analyses- e.g. subgroup analyses, sensitivity analyses | Subgroup analyses by finger, joint level, and movement type reported across Tables 2-9 (p.6-10). No sensitivity analyses conducted. No interaction analyses. |
| **DISCUSSION** | | | |
| 18 | Key results | Summarise key results with reference to study objectives | Discussion – Test-retest reliability, p.10-11; Concurrent validity, p.11-12: Reliability findings summarised by joint and movement type. Highest reliability for MCP/PIP joints. Strong concurrent validity for MCP extension. Results contextualised against prior literature. |
| 19 | Limitations | Discuss limitations, including potential sources of bias, imprecision, and both direction and magnitude of any potential bias | Limitations, p.13: (1) Thumb opposition ceiling effect in healthy population prevented ICC computation. (2) MDC not determinable without clinical population. Direction and magnitude of measurement bias not quantitatively analysed. |
| 20 | Interpretation | Give a cautious overall interpretation of results considering objectives, limitations, multiplicity of analyses, and results from similar studies | Discussion p.11-12 and Clinical Implications p.12-13: Lower r for abduction/adduction contextualised against known limitations of goniometry as reference standard. Prior literature cited (refs 16-21). Cautious interpretation that lower r may reflect reference standard imprecision rather than tool error. |
| 21 | Generalisability | Discuss the generalisability (external validity) of the study results | Conclusion, p.13: Results explicitly limited to healthy adult population. Future validation in neurological populations (stroke, cerebral palsy) recommended before clinical translation. |
| **OTHER INFORMATION** | | | |
| 22 | Funding | Give the source of funding and the role of the funders for the present study and, if applicable, for the original study on which the present article is based | Funding, p.13: 'This study received no funding from public, commercial, or nonprofit agencies.' |
